# A rapid and cost-effective multiplex ARMS-PCR method for the simultaneous genotyping of the circulating SARS-CoV-2 phylogenetic clades

**DOI:** 10.1101/2020.10.08.20209692

**Authors:** Md. Tanvir Islam, A. S. M. Rubayet Ul Alam, Najmuj Sakib, Md. Shazid Hasan, Tanay Chakrovarty, Md. Tawyabur, Ovinu Kibria Islam, Hassan M. Al-Emran, Iqbal Kabir Jahid, M. Anwar Hossain

## Abstract

Tracing the globally circulating SARS-CoV-2 mutants is essential for the outbreak alerts and far-reaching epidemiological surveillance. The available technique to identify the phylogenetic clades through high-throughput sequencing is costly, time-consuming, and labor-intensive that hinders the viral genotyping in low-income countries. Here, we propose a rapid, simple and cost-effective amplification-refractory mutation system (ARMS)-based multiplex reverse-transcriptase PCR assay to identify six distinct phylogenetic clades: S, L, V, G, GH, and GR. This approach is applied on 24 COVID-19 positive samples as confirmed by CDC approved real-time PCR assay for SARS-CoV-2. Our multiplex PCR is designed in a mutually exclusive way to identify V-S and G-GH-GR clade variants separately. The pentaplex assay included all five variants and the quadruplex comprised of the triplex variants alongside either V or S clade mutations that created two separate subsets. The procedure was optimized in the primer concentration (0.2-0.6 µM) and annealing temperature (56-60°C) of PCR using 3-5 ng/µl cDNA template synthesized upon random- and oligo(dT)-primer based reverse transcription. The different primer concentration for the triplex and quadruplex adjusted to different strengths ensured an even amplification with a maximum resolution of all targeted amplicons. The targeted Sanger sequencing further confirmed the presence of the clade-featured mutations with another set of our designed primers. This multiplex ARMS-PCR assay is sample, cost-effective, and convenient that can successfully discriminate the circulating phylogenetic clades of SARS-CoV-2.

## Introduction

SARS-CoV-2 has spread across 188 countries/regions within the first six months of COVID-19 pandemic infecting more than 354 million people (Dong, Du, & Gardner, 2020). This highly infectious virus poses a single-stranded positive-sense RNA genome of nearly 30 kbp (Mousavizadeh & Ghasemi, 2020). Both synonymous and non-synonymous mutations were identified in the genomic region that code for non-structural proteins (NSP1-16), structural proteins (spike, membrane, envelope, and nucleocapsid proteins), and/or seven other accessory proteins (ORF3a, ORF6, ORF7a, ORF7b, ORF8a, ORF8b, ORF8, and ORF10) (M. R. Islam et al., 2020; Kamitani, 2020; Liu, Fung, Chong, Shukla, & Hilgenfeld, 2014; Ou et al., 2020). Researchers have demonstrated that the predominant mutations may attribute to virulence (Alam, Islam, Hasan, et al., 2020; Rahman et al., 2020; Q. Wang et al., 2020). The virus has been classified into six clades namely GH, GR, G, V, L, and S by the global initiative on sharing all influenza data (GISAID) (Shu & McCauley, 2017) by the clustered, co-evolving, and clade-featured point mutations.

The mutations at position C241T along with C3037T, C14408T (RdRp:p.P323L), and A23403G (S:p.D614G) was referred as G clade. Additional mutation to the G clade at N protein:p.RG203-204KR (GGG28881-28883AAC) and ORF3a:p.Q57H (G25563T) refers to GR and GH clade, respectively. The V clade was classified by co-evolving mutations at G11083T (NSP6:p.L37F) and G26144T (ORF3a:p.G251V) where S clade strains contain C8782T and T28144C (NS8:p.L84S) variations, respectively. The L clade strains are the original or wild version for the featured mutations of five clades (Mercatelli & Giorgi, 2020).

Previous studies showed the prevalence of phylogenetic clades were different by regions and times and were closely related to variable death-case ratio (Alam et al., 2020; Toyoshima et al 2020, nature). G clade variant was dominant in Europe (Korber et al., 2020) and USA (Brufsky, 2020) on the eve of pandemic which caused high mortality in USA. This mutation variant has gradually been circulated in Southeast Asia (Alam et al., 2020; Islam et al., 2020) and Oceania (Mercatelli & Giorgi, 2020). On the contrary GR and GH clades were emerged at the end of February 2020 and GR mutant are now the leading type that cause more than one-third of infection globally (Mercatelli & Giorgi, 2020). Therefore, it is indispensable to identify the circulating clades in a specific region. Besides, several reports speculated the occurrence of SARS-CoV-2 reinfection by phylogenetically different strains that belongs to separate clades (Li et al., 2020; To et al., 2020). The dominance of a particular viral clade over others might determine the virulence, disease severity, and infection dynamics (Alam et al., 2020). However, the implications of different clades on effective drug and vaccine development is yet to be clearly elucidated (Chellapandi & Saranya, 2020).

The identification of phylogenetic clades requires the identification of specific mutations into viral genome. This identification is performed by the whole genome sequence through NGS technique that has now scaled up the deposited sequences number in GISAID to 139,000 as of October 6, 2020. Another high-throughput NGS alternative is based on clade-based genetic barcoding that targets PCR amplicons encompassing the featured mutation as described by Guan et al. (2020). However, this state of art technique has limited access to most laboratories in low-income countries. A short-throughput and small-scale genotyping would be the Sanger based targeted sequencing approach (Alam, Islam, Rahman, Islam, & Hossain, 2020), but this is labor intensive, time-consuming, inconvenient, and difficult to perform at low cost. Therefore, we have hardly observed the worldwide distribution of circulating clades in many countries, like Afghanistan, Maldives, Iraq, Syria, Yemen, Ethiopia, Sudan, Zimbabwe, Bolivia, Paraguay, and Chile, most probably due to the lack of sequencing facilities and appropriate technical personnel to perform this state-of-the-art technique. PCR-based point mutation discriminating technique, which is also known as the amplified refractory mutation system (ARMS), has been proven to be useful in identifying subtypes or clades of other respiratory viruses previously (Brister, Barnum, Reedy, Chambers, & Pusterla, 2019; Lee, Kim, Shin, & Song, 2016; W. Wang et al., 2009). In this study, we aimed to develop and validate an ARMS-based novel multiplex-PCR to identify the clade-specific point mutations of the circulating SARS-CoV-2 clades.

## Methods and materials

### Sample collection and cDNA preparation

Nasal and oral samples were collected in the health care facilities in the south-west part of Bangladesh and sent to the genome center, Jashore University of Science and Technology, Bangladesh. RNA was extracted from those samples using nucleic acid extraction kit, Invitrogen inc. The extracted RNA was then tested for SARS CoV-2 using a commercial kit from Sansure Biotceh Co., Ltd (China). The left-over RNA were preserved at −40°C in the genome center lab. A total of 24 randomly selected SARS CoV-2 positive samples were tested for the analysis (**supplementary Table s1**). A representative SARS CoV-2 negative sample (of 5 of the negative samples randomly selected) was used as a negative control in this study (**supplementary Table s2**). cDNA was prepared for each selected sample using the GoScript™ Reverse Transcription System (Promega, USA) following the manufacturer’s protocol. In brief, primer/ RNA mix was prepared by mixing 10µl of extracted RNA with 1µl of Random primer and 1µl of Oligo(dT)_15_ primer (total volume 12µl). Then the mixture was heated at 70°C for 5 minutes, followed by immediate chilling on ice for 5 minutes and a quick spin. The mixture for reverse transcription reaction was prepared by making a cocktail of the components from GoScript™ Reverse Transcription System in a sterile 1.5ml micro centrifuge tube keeping on ice. The final reaction mix was 40μl for each cDNA synthesis reaction to be performed.

### Design and in silico validation of variant-specific (3′-SNP) multiplex primers

A set of 15 primers (**Table 1**) was designed based on the ARMS for differentiating six major clades of SARS-CoV-2: S, L, V, G, GH, and GR. We designated here the L clade strains as the wild type and others as mutants. For each clade apart from L, we selected a single representative SNP variant, including 23403 A>G (p.D614G), 25563 G>T (p.Q57H), 28882 G>A (p.R203K), 26144 G>T (p.G251V) and 28144 T>C (p.L84S) from the multiple co-evolving mutations of the clades (G, GH, GR, V and S, respectively). For example, the S clade is deviated from the L clade by two mutations: C8782T, T28144C. The ‘T’ or ‘C’ at 28144 positions was rendered as wild (L clade) or mutant type variant (indicating S clade), respectively. Details of other clade-specific mutations can be derived from the GISAID site and this literature. As established for ARMS technique, this specificity was directed towards the 3′-end of the annealed primer-template (Fig. 1). The forward or reverse type-specific primers were paired with counterpart reverse or forward primer. The amplicons were simultaneously distinguished by their molecular weight (bp) in multiplex PCR in different combinations. The positive amplification of wild type targeting primers was determined as the L type. The other types were determined based on the co-evolving mutation at respective sites.

**Table 1.**
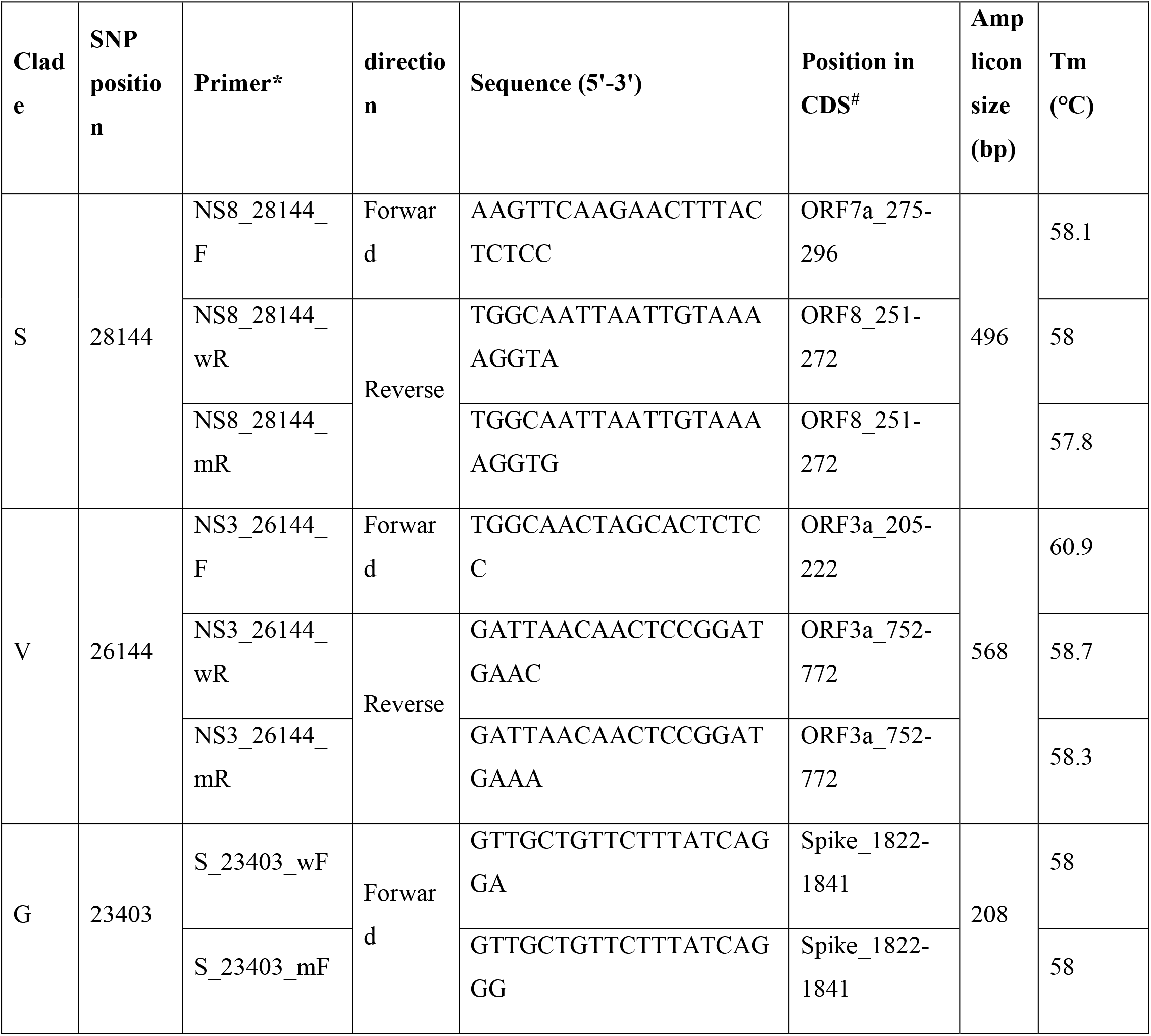
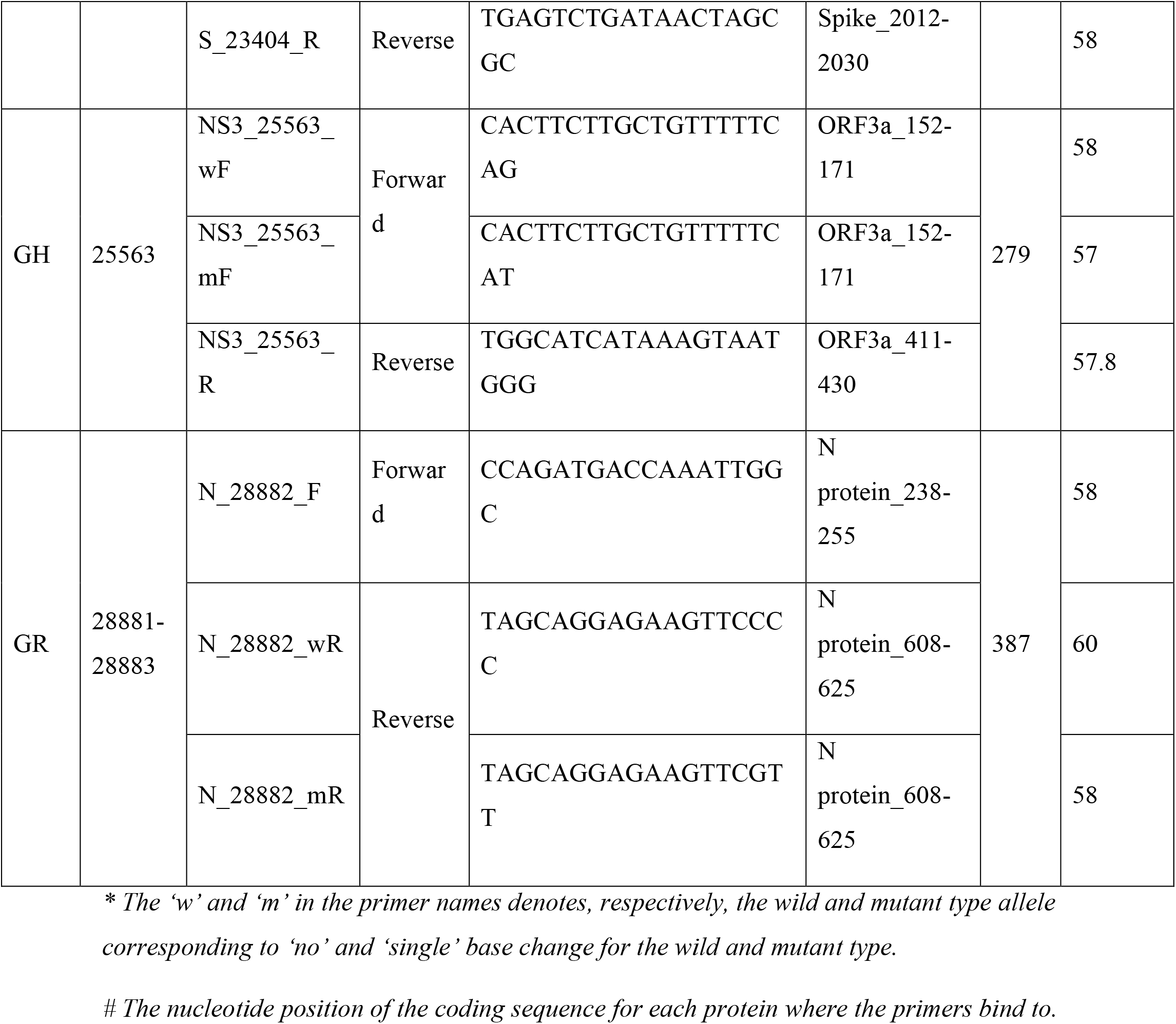
Primer sets for targeted SNP-based single- and/or multi-plex PCR.

**Figure 1.**
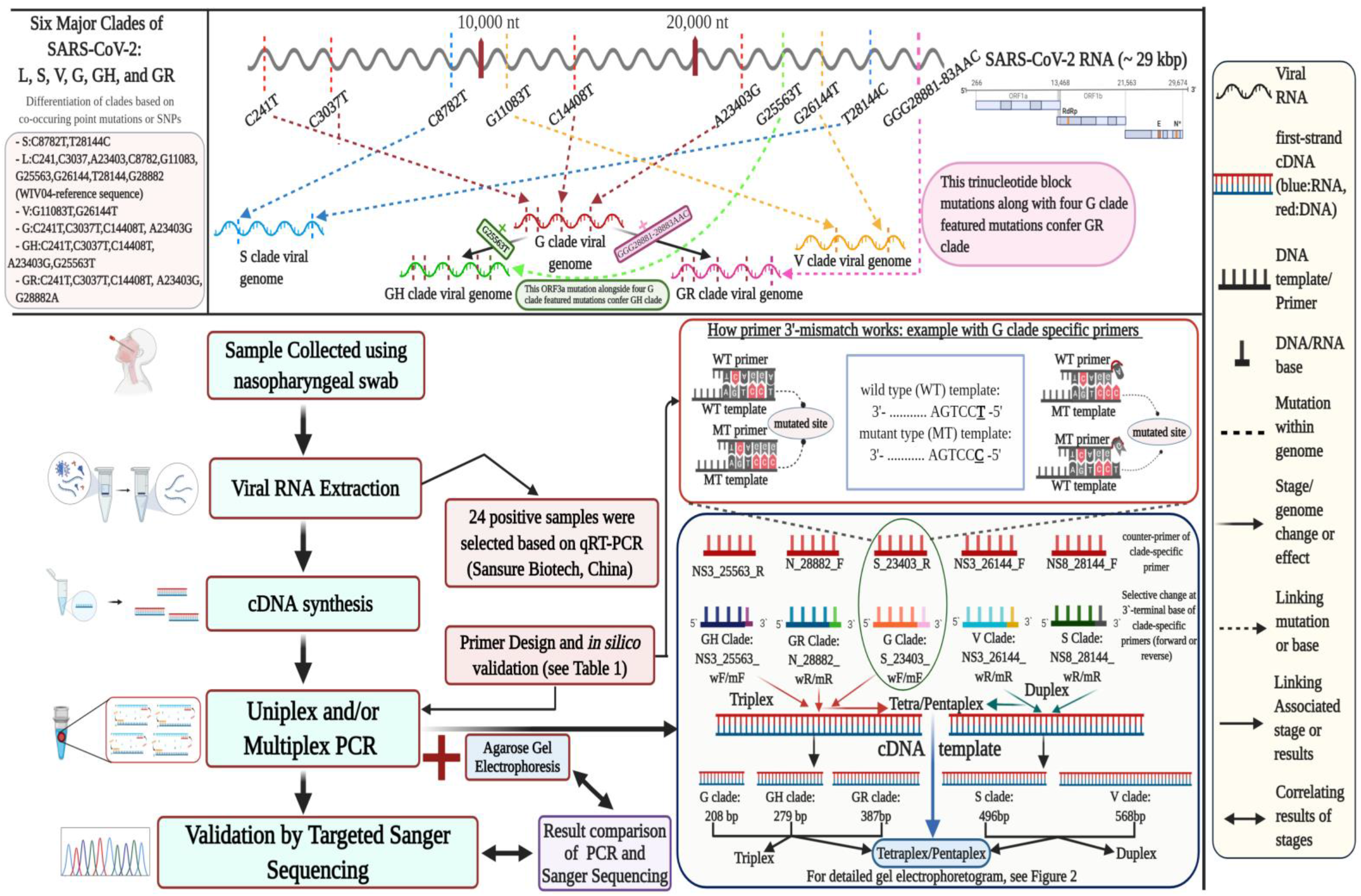
Schematic workflow of ARMS-based multiplex PCR assays for the identification of SARS-CoV-2 clades. The upper portion of the figure showed the concept of clade as described in the GISAID with a comprehensive genomic visualization. The lower segment is dedicated to the overall workflow and the primer design.

The primer sets were designed using Primer3Plus (Untergasser et al., 2012) and Primer-BLAST (Ye et al., 2012) with the following stringent parameters and standard PCR conditions: avoiding hypothetical primer dimer (self or hetero) formation with less than −9 Kcal/mol, sized 18-22 nucleotide in length, Tm of (58-60)°C, (40-60)% GC content, G/C within the last five bases, no repeat of four or more of any base, amplicon size ranging from 200 to 600 bp, and avoiding hairpin loop structure. The primer specificity against SARS-CoV-2 and other organisms was checked by the Primer-BLAST. We performed the *in silico* PCR with the primers in the UCSC genome browser (https://genome.ucsc.edu/). Finally, the primer set was synthesized from the IDT company (https://www.idtdna.com/).

### Standardization of annealing temperature for Single-variant specific PCRs

A gradient PCR (SimpliAmp Thermal Cycler, Applied Biosystems, USA) was performed for each of the variant separately with freshly prepared cDNA template to standardize the annealing temperature. Couple of distinct tubes was prepared for each of the variant using the respective primer pairs to differentiate between the wild type and the mutant. The PCR was carried out in 10µl reaction volume containing 3-5 ng/µl DNA, 5µl master mixture (GoTaq® G2 Green Master; Promega, USA), 0.2µM of each forward and reverse primer and 2.8 µl nuclease free water. The thermocycling conditions were as follows: initial denaturation at 95°C for 1 min followed by 30 cycles at 95°C for 30s, annealing at a range of 55-65°C for 30s and 72°C for 30s followed by a final extension at 72°C for 5min. The PCR products were electrophoresed on a 1% (w/v) agarose gel stained with ethidium bromide (UltraPure™ Ethidium Bromide, 10 mg/mL; Thermo Fisher, USA) and visualized using a gel documentation system (Bio-Rad, USA).

### Multiplex PCR assays for simultaneous identification of the variants

Four sets (duplex, triplex, quadruplex and pentaplex) of multiple variant-specific reactions were arranged for simultaneous detection of a clade. A duplex PCR was performed by using a mix of 26144 G>T(p.G251V) and 28144 T>C (p.L84S) variant specific primer pairs viz. NS3_26144_F-NS3_26144_wR (wild type)/NS3_26144_F-NS3_26144_mR (mutant) and NS8_28144_F-NS8_28144_wR (wild type)/ NS8_28144_F-NS8_28144_mR (mutant), respectively while mixing for wild types and the mutants in separate PCR tubes. A triplex PCR assay was performed by using a blend of primer pairs viz. S_23403_wF-S_23403_R (wild type primers)/ S_23403_mF-S_23403_R (mutant primers), NS3_25563_w1F-NS3_25563_1R (wild type primers)/ NS3_25563_m1F-NS3_25563_1R (mutant), and N_28882_F-N_28882_wR (wild type)/ N_28882_F-N_28882_mR specific to 23403 A>G (p.D614G), 25563 G>T (p.Q57H) and 28882 G>A (p.R203K) SNP variants respectively. Quadruplex and pentaplex PCR assays were further performed in similar manner. The pentaplex consisted the mix for all five SNP variants, On the other hand, the quadruplex contained the variants of triplex plus either 26144 G>T (p.G251V) or 28144 T>C (p.L84S), thus making two different subsets. Among the 24 samples, one was used as a representative of all sets of multiplexes and reproducibility (described in more detail below) was checked over the rest of the samples.

### Validation of multiplex PCR assays

The multiplex PCR assays were performed over all the 24 positive samples. To validate the reliability of the assays, another five pairs of primer set (**Table 2**) were designed for the clades keeping the probable mutation points within the middle of the amplicons by using Primer3Plus (Untergasser et al., 2012) and Primer-BLAST (Ye et al., 2012). Above mentioned parameter settings were followed to design those (except for the amplicon size ranging from 200-400bp and Tm of 52-54°C). Amplicons were subjected to Sanger sequencing using BigDye™ Terminator v3.1 Cycle Sequencing Kit (ThermoFisher Scientific) to confirm the specific clades (wild type/mutant). The commercial kit protocol was optimized to reduce the cost because of the high cost of this BigDye Terminator reagent (Platt, Woodhall, & George, 2007). 1.0μL (per 10 10μL reaction) undiluted BigDye Terminator v3.1 Ready Reaction mix was used instead of 4μL mentioned in commercial kit protocol. Along with the 1.0 μL BigDye Terminator v3.1 Ready Reaction mix, 1.75 μL 5x sequencing buffer, 1μL primer, 2μL template DNA and 4.25μL nuclease free water was added (to make the final reaction volume 10μL). The cycle sequencing PCR condition was setup accordingly to the kit protocol. The sequences were aligned with and verified by the reference sequence (NC_045512.2_SARS-CoV-2_Wuhan-Hu-1 complete genome) using Molecular Evolutionary Genetics Analysis (MEGA X) software (Kumar, Stecher, Li, Knyaz, & Tamura, 2018). For the most part, the cost of the processes were optimized and compared to our in-house NGS system (Ion torrent; ThermoFisher Scientific, USA).

**Table 2.**
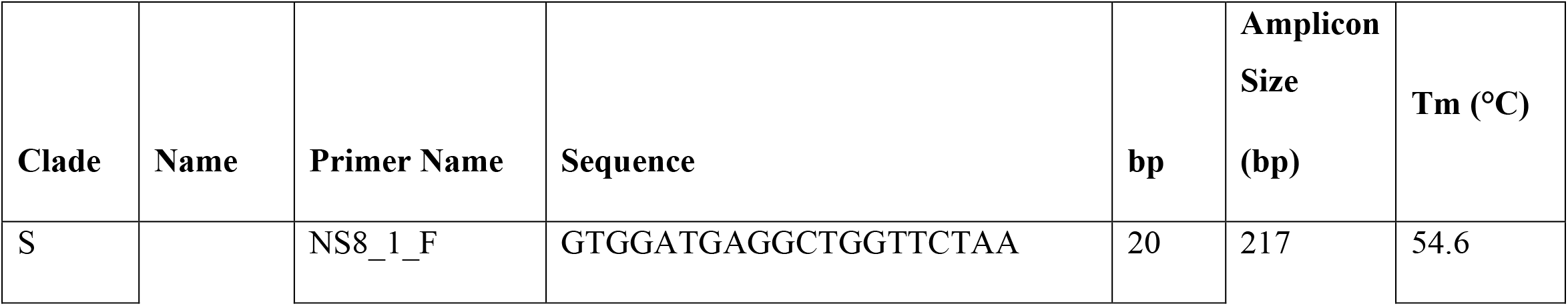
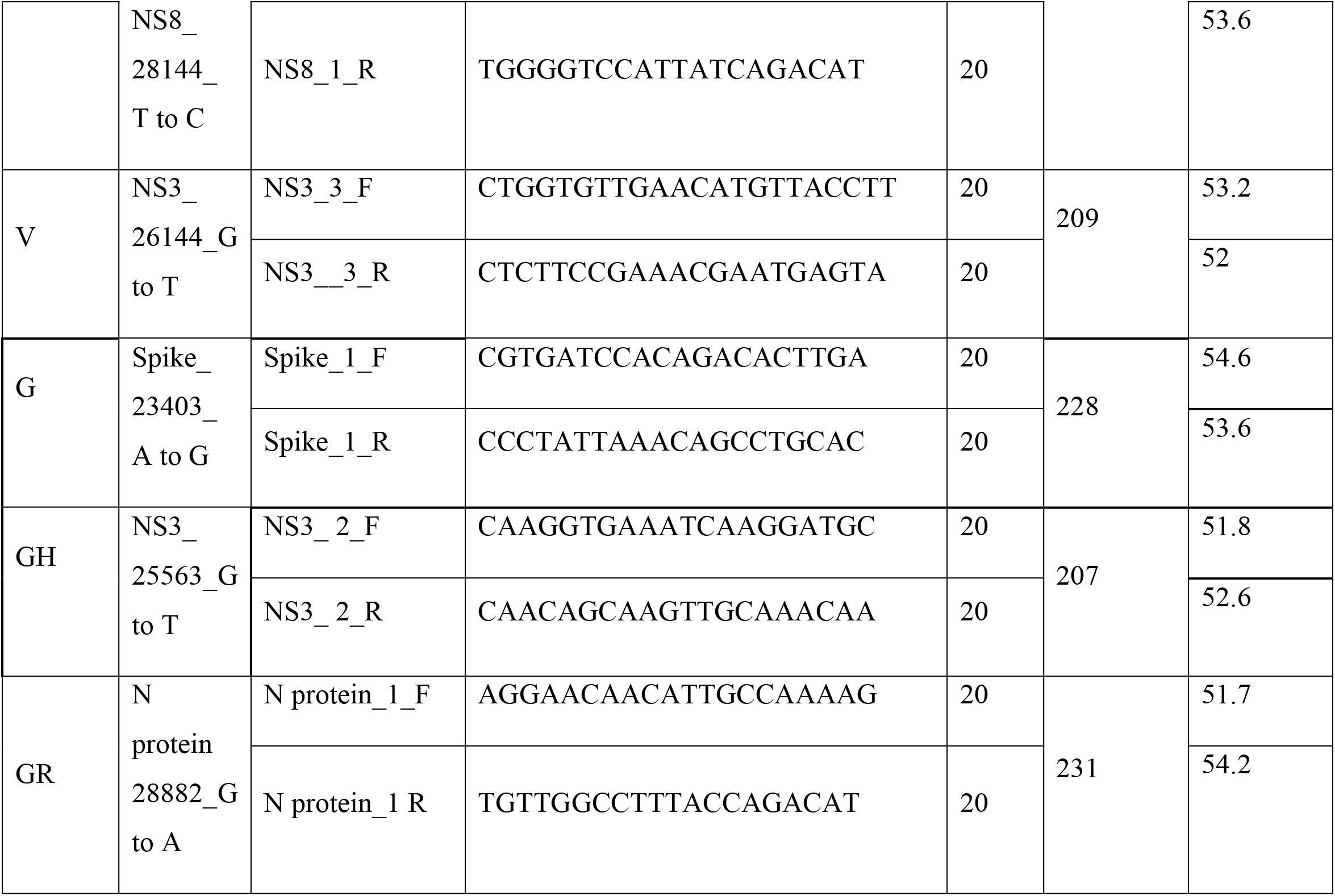
List of primers for targeted Sanger sequencing to validate the multiplex assays.

### Prediction of Primer dimer formation and RNA secondary structure

We carried out RNA secondary structure prediction of the ORF3a or NS3 RNA using the Mfold web server (Zuker, 2003). The full NS3 sequence was extracted from the SARS-CoV-2 reference sequence of NCBI GenBank. The default parameters were used in generating structure. Besides, we used oligoanalyzer v3.1 of integrated DNA technologies (IDT) to examine possible primer duplexes and calculate the primer-dimer (both self and hetero) formation energy in case of the primers, NS3_26144_F, NS3_26144_wR, and NS3_26144_mR, targeting ORF3a multiplexed amplicons. The possibility of dimers and energy values for the targeted primers were checked against other twelve primer sets.

## Results

### Identification of the clades by single-variant specific PCRs

ARMS-based PCRs for single SNP variants (singleplex PCRs) were optimized at an annealing temperature of 57°C. The primer pairs for the variants 23403 A>G (p.D614G) and 28882 G>A (p.R203K) amplified the related mutant bands (208bp and 387bp, respectively). On the other hand, the primer pairs designed to denote the variants 25563 G>T (p.Q57H), 26144 G>T(p.G251V) and 28144 T>C (p.L84S) had wild type amplicons (279bp, 568bp and 496bp, respectively) (**Fig.2 IA-IE**). Hence, the single-variant specific PCRs were able to identify the SARS-CoV-2 positive sample containing GR-clade of the virus.

**Figure 2.**
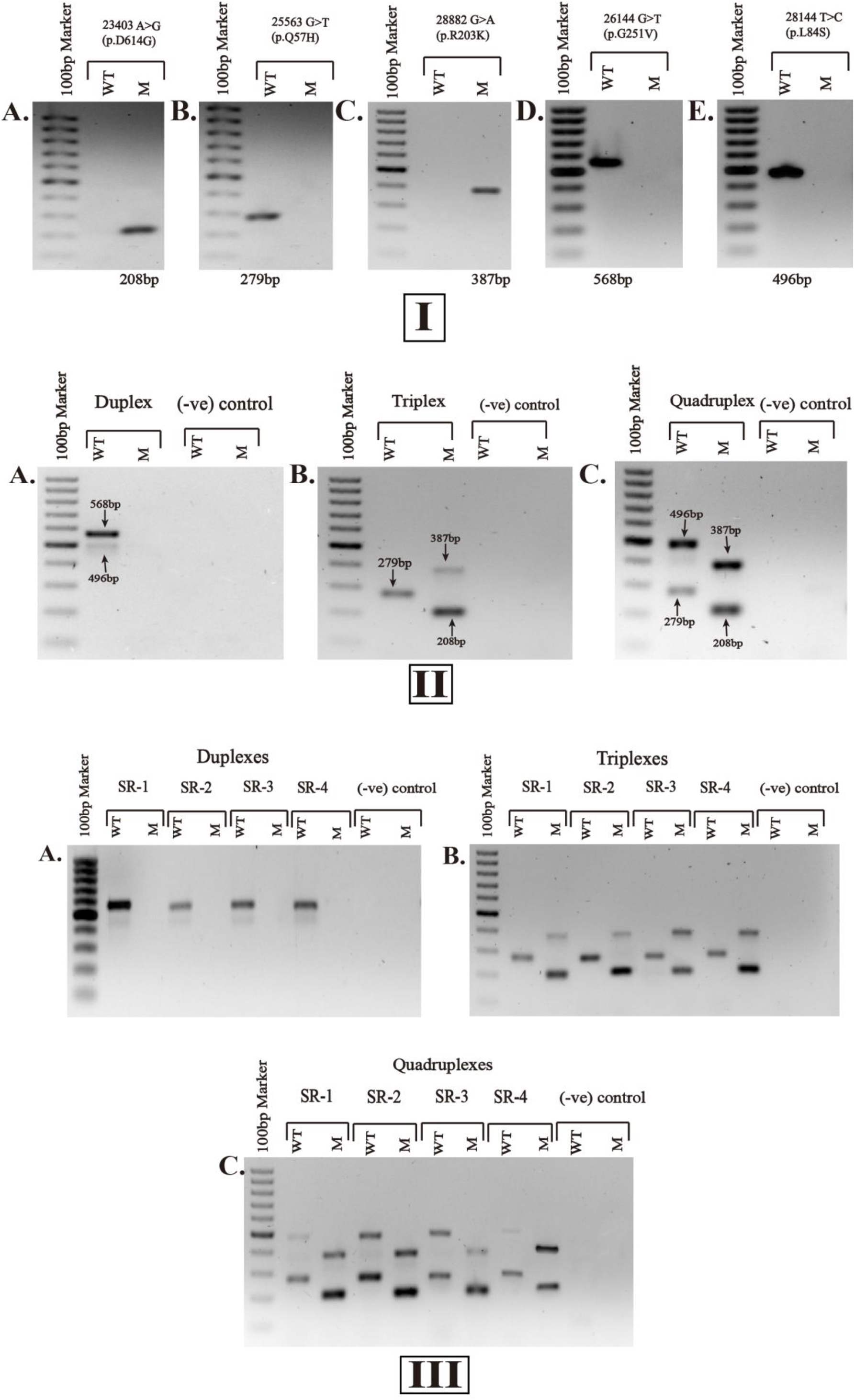
Strategy and validation of ARMS-based multiplex PCR assays. **(I)** Single-variant specific PCRs with the respective amplicons in bp at the bottom of each gel image, showing the variants. **(II)** Multiplex PCR assays, containing PCR products for different clade combinations: duplex for 26144 G>T (p.G251V) and, 28144 T>C (p.L84S) variants at an annealing temperature of 60°C; triplex for the variants 23403 A>G (p.D614G), 25563 G>T (p.Q57H) and, 28882 G>A (p.R203K) at 57°C; quadruplex for the variants 23403 A>G (p.D614G), 25563 G>T (p.Q57H), 28882 G>A (p.R203K) and, 28144C T>C (p.L84S) at 56°C. SARS-CoV-2 positive sample, GC 46.003 was used as a representative to perform the PCRs for (I) and (II). **(III)** Validation of the multiplex PCR assays, containing the identical settings for duplex, triplex and quadruplex PCRs with GC 44.201 (denoted as SR-1), GC 88.025 (denoted SR-2), GC 90.175 (denoted SR-3) and GC 92.172 (denoted as SR-4) as representatives to display the reproducibility of the assays. SARS-CoV-2 negative sample, GC 116.09 was used as (-ve) control for the comparison. ‘GC’ indicates the Genome Center identification number generated at the Genome Center of Jashore University of Science and Technology for COVID-19 suspected patients. WT, wild type band; M, Mutant band. 100-1000bp marker was used in the first lane of the 1% agarose gels. The primers are listed in Table 1.

### Optimization of multiplex PCR assays

The singleplex PCRs showed successful annealing at 57°C, however, temperatures for the duplex, triplex and quadruplex assays were needed to be further optimized to 60°C, 57°C and 56°C, respectively (**Fig.2 IIA-IIE**). The primer concentrations for duplex were similar (0.2µM of each primer pair for both the SNP variants) to the single-variant specific PCRs, they were adjusted to different strengths for the triplex (for both forward and reverse primers: 0.2µM for 23403 A>G (p.D614G), 0.3µM for 25563 G>T (p.Q57H), and 0.4µM for 28882 G>A (p.R203K)) and the quadruplex (for both forward and reverse primers: 0.4µM for 23403 A>G (p.D614G), 0.3µM for 25563 G>T (p.Q57H), 0.6µM for 28882 G>A (p.R203K) and 0.2µM for 28144 T>C (p.L84S)) to get the maximum possible resolution. The duplex PCR assay simultaneously amplified the desired wild type bands of 568bp and 496bp; the band for 28144 T>C (p.L84S) was faint in the duplex setting comparing to the single run of the variant described before. Besides, the triplex PCR also amplified the products as expected (i.e., 208bp, 279bp and 387bp). One of the subsets of quadruplex that contained the variants of triplex plus 28144C T>C (p.L84S) was able to distinguish the desired bands individually. However, quadruplex (that had 26144 G>T (p.G251V)) and pentaplex arrangement could not discriminate the bands between wild types and mutants (**supplementary Fig.s1**).

### Validation of multiplex PCR assays

All the 24 positive samples confirmed the test reproducibility of the assays; four of them excluding the one used before for multiplex assays were taken as representative to display the reproducibility in this article (**Fig.2 IIIA-IIIC**). In this study, only GR clade was found in all positive samples tested. The homology of the nucleotide sequences for the PCR products showed more than 99% identity with the respective positions of the clades that validated the assays (**supplementary Fig.s2**). Accession IDs to the submitted sequences for one positive sample as an archetype are available in GISAID EpiFlu™ database (EPI_ISL_548260, EPI_ISL_561630, EPI_ISL_561375, EPI_ISL_561376 and EPI_ISL_561377).

## Discussion

This study proposes a simple and exclusive ARMS-based SNP-discriminating method using conventional PCR to establish multiplex-assays in detecting SARS-CoV-2 mutation clades. This concept was adopted from the other studies applied to identify the genetic profile of respiratory or gastrointestinal coronaviruses of pigs, human cancer risk related SNPs, virus that causes systemic infection in canines, the resistance profile of a bacteria etc. (Chulakasian et al., 2010; Lai, Welter, & Welter, 1995; Shi et al., 2013; C. Zhang et al., 2013). This study designed point-mutation specific primers to detect the six different SARS-CoV-2 clades as described by GISAID. The clade-based discrimination during COVID-19 pandemic was exceedingly important because the prevalence of SARS-CoV-2 clades were varied by regions and times, and were closely related to variable death-case ratio (Alam et al., 2020; Toyoshima et al 2020, nature). G clade variant was dominant in Europe (Korber et al., 2020) and USA (Brufsky, 2020) on the beginning of the pandemic that caused high mortality in USA. This mutation variant was gradually circulating in Southeast Asia (Alam, Islam, Hasan, et al., 2020; O. K. Islam et al., 2020) and Oceania (Mercatelli & Giorgi, 2020). Among the two derivative of G clade (GR and GH) that emerged at the end of February 2020, GR mutant are now the leading type that cause more than one-third of infection globally (Mercatelli & Giorgi, 2020).

In this study, we attempted to validate two sets of multiplex PCR covering G, GH and GR in the first set, and V and S in the second set. Based on the available data of clade prevalence we propose to run the first set of multiplex PCR in the beginning that can confirm the most three prevalent clades (Alam, Islam, Hasan, et al., 2020). Our attempt for pentaplex and/or the quadruplex (that included the SNP variant 26144 G>T (p.G251V)) was unsuccessful, where the template regarding the variant 26144 G>T (p.G251V) did not amplify, possibly due to primer-dimer formation with higher thermodynamic stability than other variant-specific primer sets. The forward primer could bind to the N_28882_mR primer with a G value of <-7 Kcal/mol, but can make longer products ∼40 bp. In case of reverse primers that target mutation, only NS3_26144_wR would form a self-dimer with high free energy (−12.9 Kcal/mol). These homo- and hetero-dimer formation would make more primer duplex and might reduce the chance of effective pentaplex PCR. Another possibility could be that the NS3 binding region of the primers (205-222 and 752-772) has resided in a stable stem site hindered the effective annealing. The RNA structure showed the complex stem-loop region and open sites as well. Our targeted primer binding sites for the variant 26144 G>T (p.G251V) were within the stem-loop region whereas the primer annealing sites for 25563 G>T (p.Q57H) variant reside within the open region of the template. Here we assume, the complex structural configuration of NS3 may block the PCR reaction during a competitive multiplexing (**supplementary Fig.s3**). A longer RNA denaturation step during cDNA synthesis and more stringent cycle denaturation of cDNA template might solve this issue. However, it could damage low-concentrated, sample-extracted viral RNA and inhibit amplification of other clade-specific templates by affecting overall optimized multiplex condition.

The advantage of our ARMS-based multiplex assays is rapid. The turnaround time for our designed assay would range from approximately 3-4 hours for 96 samples. The NGS and Sanger methods, on the other hand, had a turnaround time of more than 24 hours and 10-12 hours, respectively (Tsuchihashi & Dracopoli, 2002; J. Zhang et al., 2020). An ARMS based multiplex PCR assay similar to the current study would render a more convenient way to detect clade specific mutation (SNPs) due to the process being faster and cost effective (Ahlawat, Sharma, Maitra, Roy, & Tantia, 2014). The cost of the assay for a single reaction was $7 per run (the cost includes import Tax and VAT etc. for Bangladesh) that is much less than targeted and whole-genome based NGS methods in identifying the clades. The cost will be further reduced if an optimized one-step PCR system is used and we are currently working on it to cut the overall cost down to less than $2. Thus, our method can overcome a serious limitation to effectively identify viral clades with a prospective broader application. The requirement of technical skill would also be low for this assay wherein the training of personnel is a minimal requirement and interpretation of results is generic (Syrmis et al., 2004; L. Wang et al., 2011). Besides, the presence of the template as well as their quantity and quality are determined at the same time. The false-negative result for the absence of a template can also be determined in a facile manner (Edwards & Gibbs, 1994). In general, mutating the primer at its 3’prime end makes it refractory to the ‘wild type template’ whereas the absence of mutation in the primer is retractable to the ‘mutant template’ amending a reliable technique over sequencing (Chulakasian et al., 2010). On the other hand, next generation sequencing technology such as whole genome sequencing (WGS) or metagenomics approach can generate millions of high-throughput data that enabled researchers to unroll new dimensions in the field of genome sequencing applications (El-Metwally, Hamza, Zakaria, & Helmy, 2013). The lack of technical personnel to analyze NGS data is also a reason to prefer alternative approach other than NGS technology in low-income countries. Therefore, the ARMS technology with the conventional multiplex PCR methods in identifying the clades would be more applicable in low and minimum resource settings.

## Conclusion

Our assay can enhance the identification of genotypic variants of SARS-CoV-2 worldwide, especially in low-resource settings where NGS and Sanger sequencing techniques are difficult to reach out. This rapid barcoding method may assist to reveal disease epidemiology, patient management and protein-based drug designing, and also contribute to modify future national policy, and vaccine development. A more cost-effective one-step procedure based on modified tetra ARMS assay (T-ARMS) is under development by our group that will considerably reduce the labor and cost further.

## Data Availability

Data can be checked from the GISAID EpiFlu database and the accession numbers are EPI_ISL_548260, EPI_ISL_561630, EPI_ISL_561375, EPI_ISL_561376 and EPI_ISL_561377.

## Acknowledgments

We acknowledge GISAID for sharing the sequence data and IDT for giving the opportunity to use the tools for validating primers *in silico*. We also acknowledge Ministry of Health and family Welfare, Bangladesh for giving us the permission for SARS CoV-2 diagnosis.

## Authors’ contribution

OKI, ARA, and IKJ conceived and designed the study outline. MTI and MSH setup initial experiments. MTI established the methodology with the assistance of NS, TC and MT. MTI, NS processed the experimental data, interpreted the results and generated the figures. ARA designed the primers for multiplex ARMS-PCR and MSH designed the primers for targeted SNP positions. ARA performed bioinformatics and *in silico* primer validation. MSH analyzed sequence data. NS, ARA, and MSH wrote the manuscript in consultation with OKI, HMA, IKJ and MTI. HMA and IKJ further reviewed and edited the manuscript extensively. IKJ and MAH supervised the project and edited the manuscript. Final approval was provided by all authors.

## Availability of data and materials

Data can be checked from the GISAID EpiFlu™ database and the accession numbers are EPI_ISL_548260, EPI_ISL_561630, EPI_ISL_561375, EPI_ISL_561376 and EPI_ISL_561377.

## Ethics approval and consent to participate

This study was approved by the ethical review committee of Jashore University of Science and Technology (Ref: ERC/FBS/JUST/2020-45, Date: 06/10/2020). The samples have been used from the routine diagnosis.

## Consent for publication

Not applicable.

---

**Figure s1.**
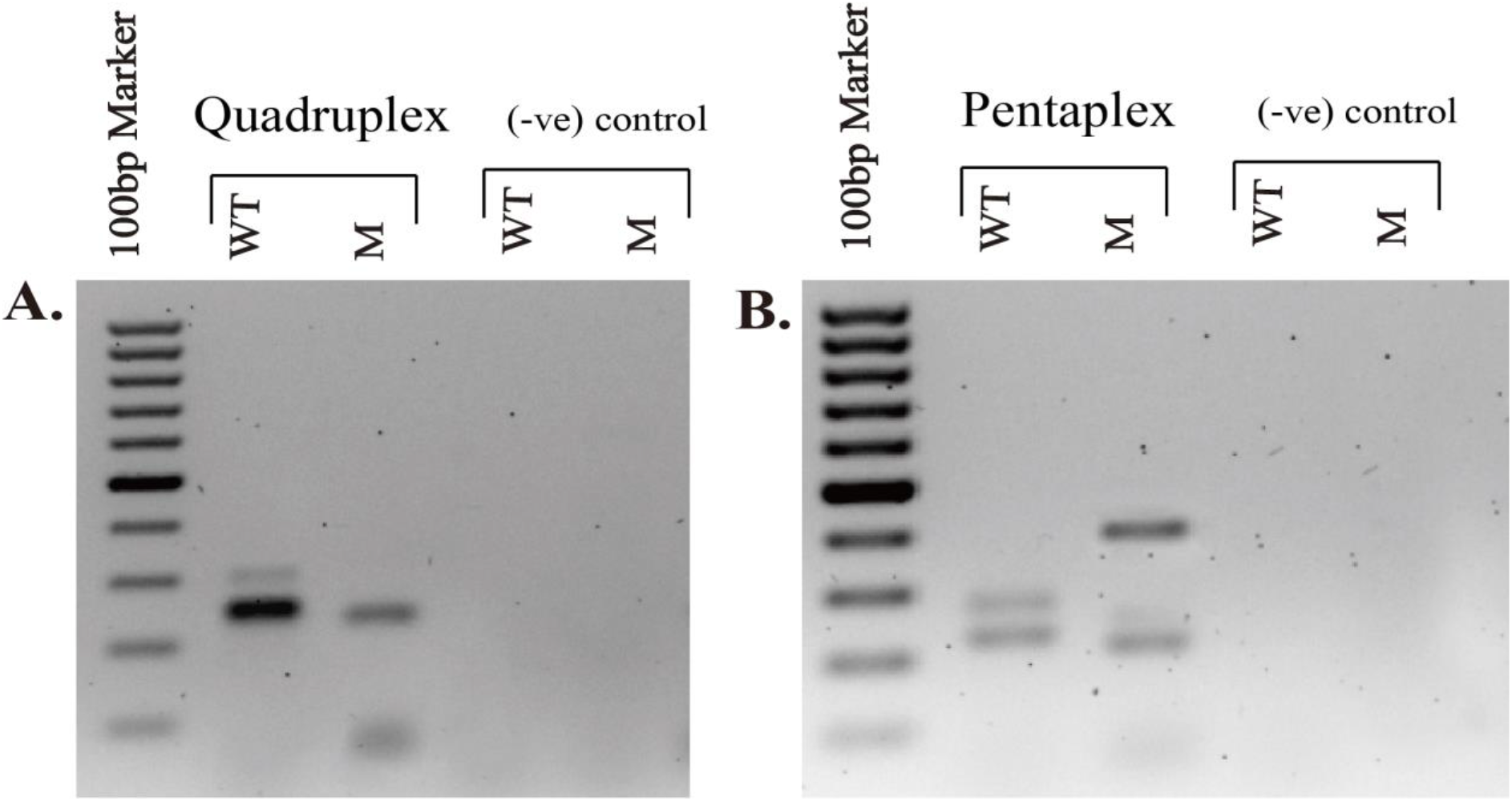
Failed attempts in generating optimized quadruplex and pentaplex assays. No optimization or adjustment could discriminate the desired location of the bands in quadruplex (A) and in the pentaplex (B). Quadruplex comprises the SNP variants 23403 A>G (p.D614G), 25563 G>T (p.Q57H), 28882 G>A (p.R203K) and, 26144 G>T (p.G251V). Pentaplex includes the SNP variants 23403 A>G (p.D614G), 25563 G>T (p.Q57H), 28882 G>A (p.R203K), 28144C T>C (p.L84S) and 26144 G>T (p.G251V). SARS-CoV-2 positive sample, GC 46.003 was used as a representative to perform the PCRs. SARS-CoV-2 negative sample, GC 116.09 was used as (-ve) control for the comparison. ‘GC’ indicates the Genome Center identification number generated at the Genome Center of Jashore University of Science and Technology for COVID-19 suspected patients. WT, wild type band; M, Mutant band. 100-1000bp marker was used in the first lane of the 1% agarose gels. The primers are listed in Table 1.

**Figure s2.**
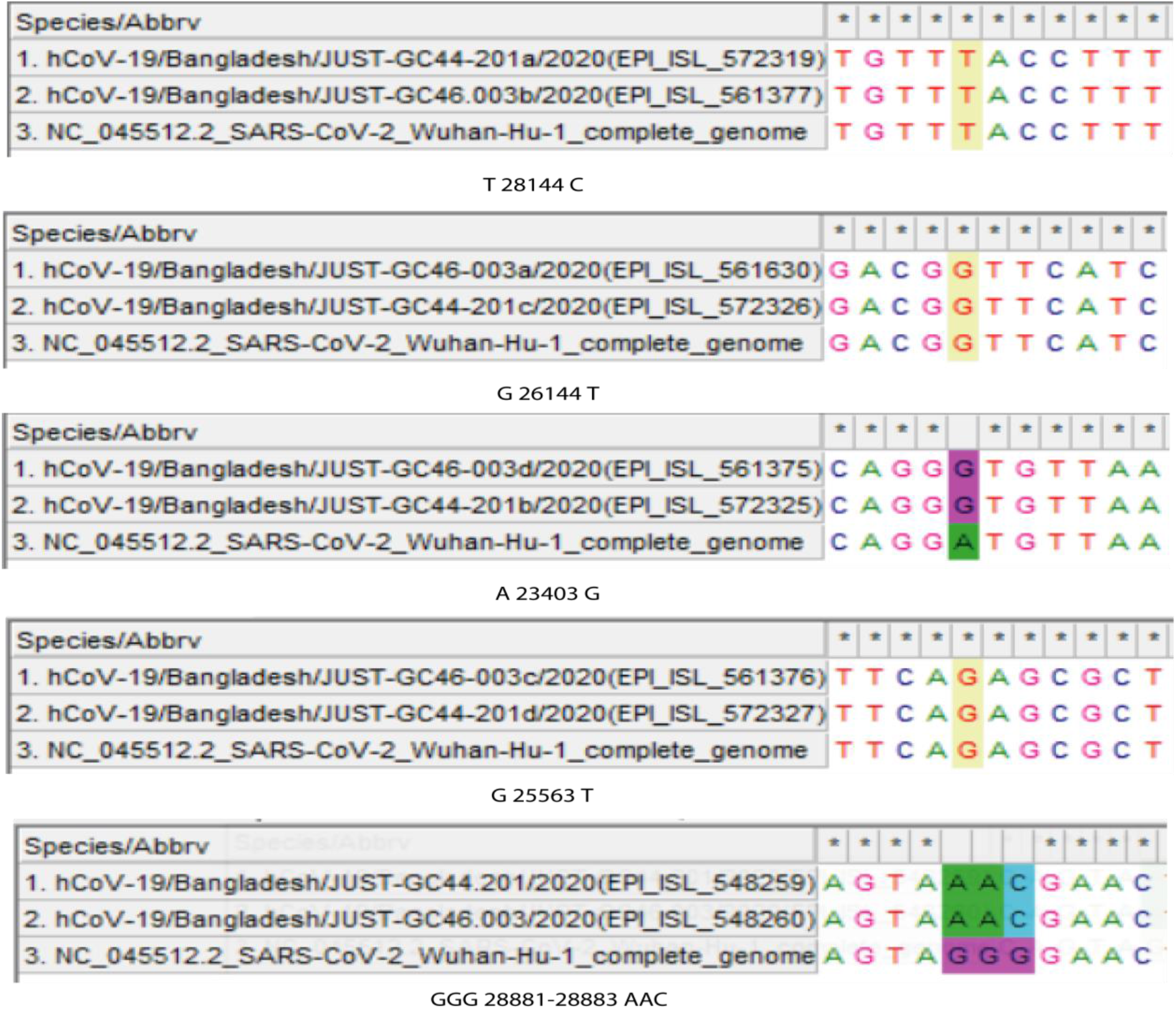
Sanger sequencing of variant-specific mutation (SNPs) positions to detect the clade of SARS-CoV-2. Alignment of the partial sequences with the reference sequence (NC_045512.2_SARS-CoV-2_Wuhan-Hu-1_complete_genome) showed that both the GC44.201and GC46.003 samples are GR Clade viruses. ‘GC’ indicates the Genome Center identification number generated at the Genome Center of Jashore University of Science and Technology for COVID-19 suspected patients.

**Figure s3.**
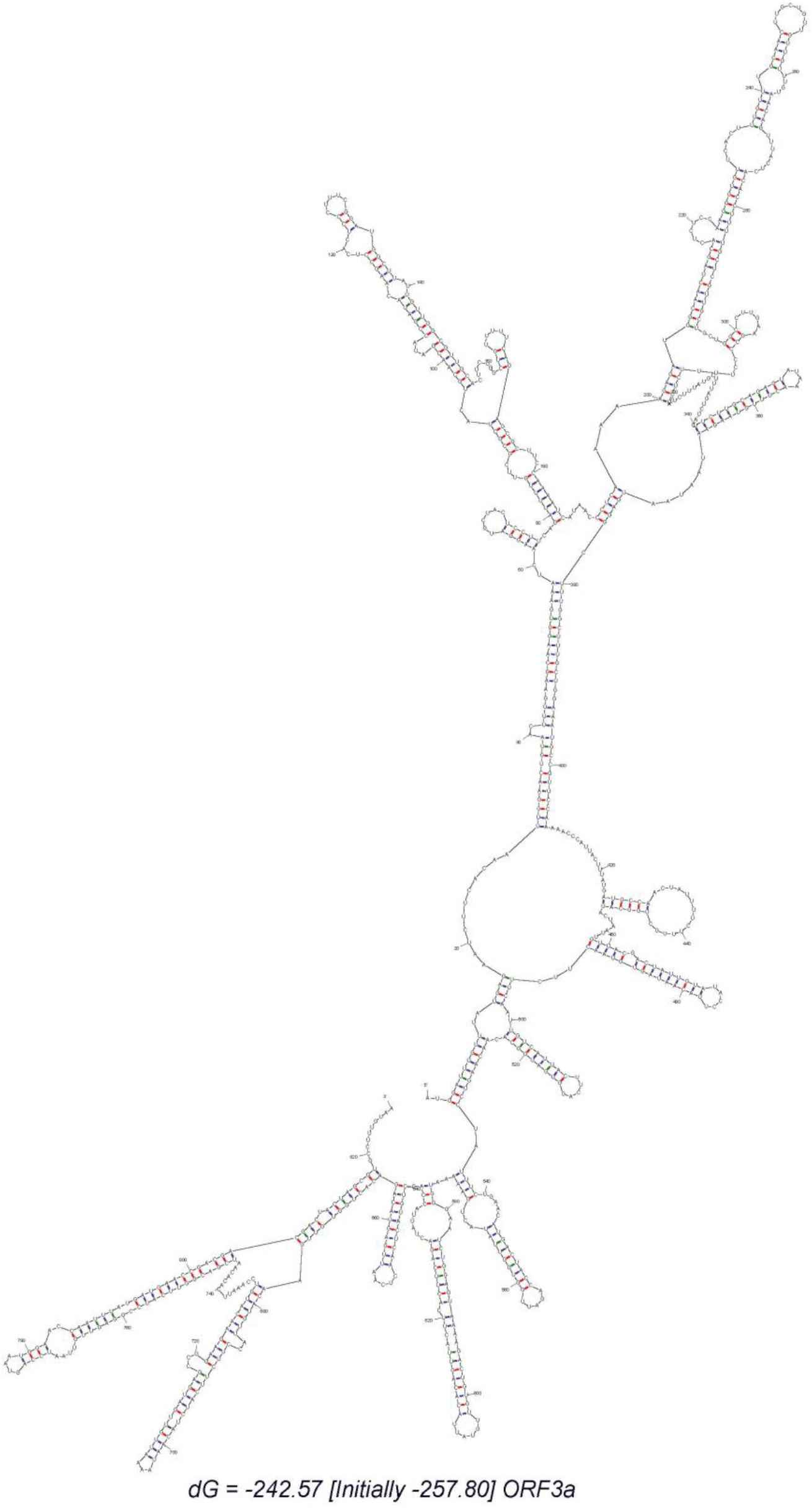
The secondary RNA structure of NS3 of ORF3a raw template. The interval is 20 nucleotides. The free energy is −242.57 Kcal/mol.

**Table s1.** List of SARS-CoV-2 positive samples examined at the Genome Center of Jashore University of Science and Technology.

|  | **Sample/**<br>**Specimen ID\*** | **District** | **Date of Sample**<br>**Collection (dd-**<br>**mm-yyyy)** | **Date of Test**<br>**(dd-mm-**<br>**yyyy)** | **Result** |
| --- | --- | --- | --- | --- | --- |
| 1 | GC36.122 | Jashore | 7/6/2020 | 8/6/2020 | Positive |
| 2 | GC37.01 | Jashore | 8/6/2020 | 8/6/2020 | Positive |
| 3 | GC42.44 | Jashore | 13/06/2020 | 14/06/2020 | Positive |
| 4 | GC43.374 | Jashore | 14/6/2020 | 16/06/2020 | Positive |
| 5 | GC44.201 | Sathkhira | 15/06/2020 | 18/06/2020 | Positive |
| 6 | GC46.003 | Jashore | 17/06/2020 | 19/06/2020 | Positive |
| 7 | GC46.026 | Jashore | 17/06/2020 | 19/06/2020 | Positive |
| 8 | GC48.152 | Bagerhat | 19/06/2020 | 20/06/2020 | Positive |
| 9 | GC48.159 | Bagerhat | 19/06/2020 | 20/06/2020 | Positive |
| 10 | GC51.183 | Jashore | 22/06/2020 | 23/06/2020 | Positive |
| 11 | GC53.091 | Satkhira | 24/06/2020 | 25/06.2020 | Positive |
| 12 | GC86.011 | Jashore | 03/08/2020 | 04/08/2020 | Positive |
| 13 | GC86.020 | Jashore | 03/08/2020 | 04/08/2020 | Positive |
| 14 | GC87.025 | Jashore | 4/08/2020 | 5/08/2020 | Positive |
| 15 | GC88.005 | Magura | 5/08/2020 | 6/08/2020 | Positive |
| 16 | GC88.131 | Jashore | 5/08/2020 | 6/08/2020 | Positive |
| 17 | GC88.222 | Jashore | 5/08/2020 | 6/08/2020 | Positive |
| 18 | GC88.025 | Magura | 5/08/2020 | 6/08/2020 | Positive |
| 19 | GC88.056 | Narail | 5/08/2020 | 6/08/2020 | Positive |
| 20 | GC91.124 | Jashore | 8/08/2020 | 9/08/2020 | Positive |
| 21 | GC90.175 | Narail | 7/08/2020 | 8/08/2020 | Positive |
| 22 | GC92.172 | Jashore | 9/08/2020 | 10/08/2020 | Positive |
| 23 | GC52.200 | Bagerhat | 23/06/2020 | 24/06/2020 | Positive |
| 24 | GC53.110 | Magura | 24/06/2020 | 25/06/2020 | Positive |

*\*‘GC’ indicates the Genome Center identification number generated at the Genome Center of Jashore University of Science and Technology for COVID-19 suspected patients.*

**Table s2:**
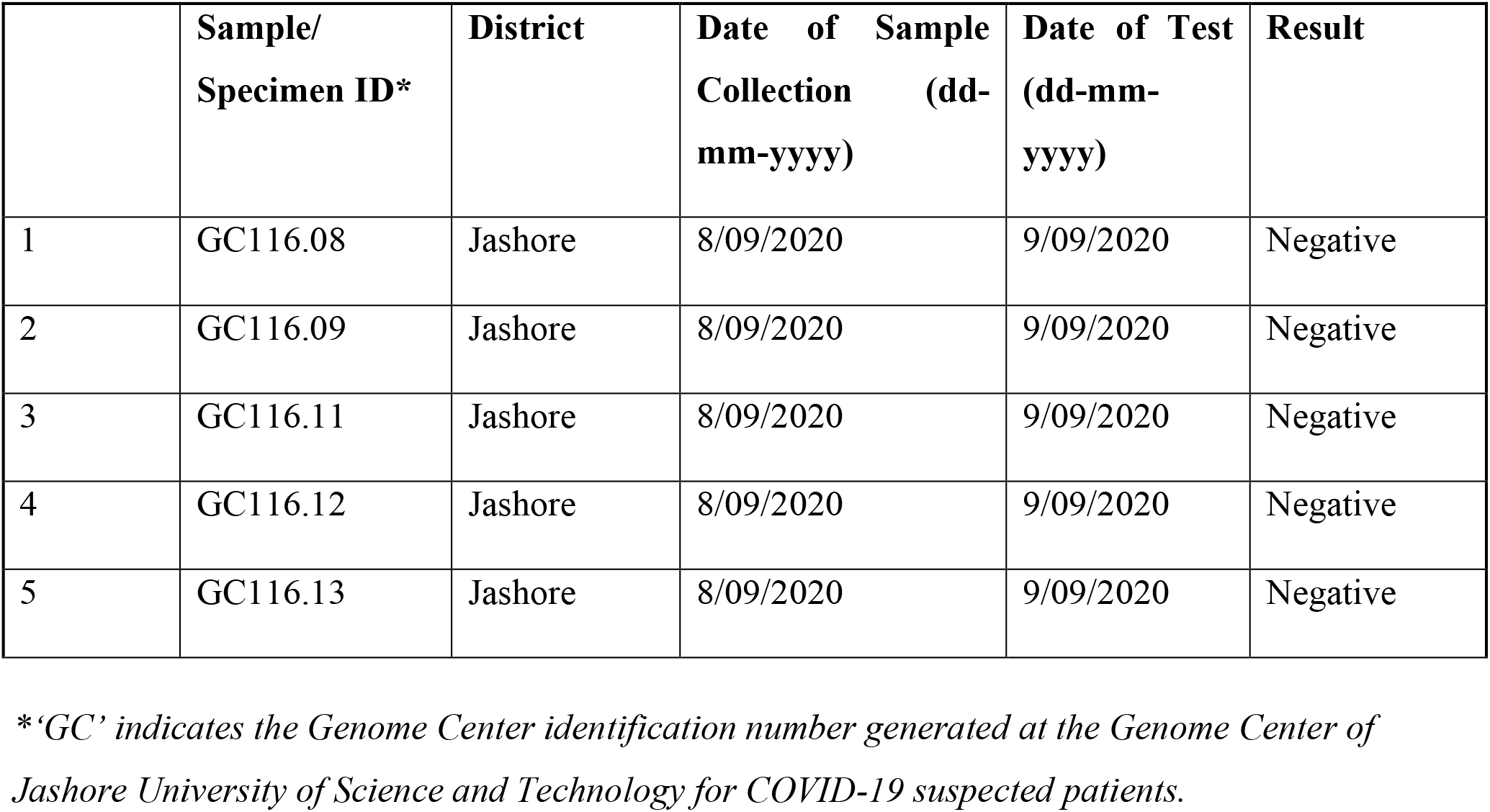
List of SARS-CoV-2 negative samples examined at the Genome Center of Jashore University of Science and Technology.

|  | Sample/<br>Specimen ID* | District | Date of Sample<br>Collection (dd-mm-yyyy) | Date of Test<br>(dd-mm-yyyy) | Result |
| --- | --- | --- | --- | --- | --- |
| 1 | GC116.08 | Jashore | 8/09/2020 | 9/09/2020 | Negative |
| 2 | GC116.09 | Jashore | 8/09/2020 | 9/09/2020 | Negative |
| 3 | GC116.11 | Jashore | 8/09/2020 | 9/09/2020 | Negative |
| 4 | GC116.12 | Jashore | 8/09/2020 | 9/09/2020 | Negative |
| 5 | GC116.13 | Jashore | 8/09/2020 | 9/09/2020 | Negative |

*\*‘GC’ indicates the Genome Center identification number generated at the Genome Center of Jashore University of Science and Technology for COVID-19 suspected patients.*

